# Effect of Glucagon-Like Peptide-1 Deprescription Following Nutrition Therapy via Telemedicine on Glycemia and Body Weight among People with Type 2 Diabetes in a Real-World Setting: A Propensity Score Matched Cohort Study

**DOI:** 10.1101/2023.06.18.23291518

**Authors:** Amy L. McKenzie, Shaminie J. Athinarayanan

## Abstract

**Introduction:** Recent clinical trials demonstrate that glucagon-like peptide-1 receptor agonists (GLP-1) elicit substantial reductions in glycemia and body weight in people with type 2 diabetes and obesity but must be continued indefinitely to maintain clinical improvements. Given the high cost and poor real-world persistence of GLP-1, an effective maintenance therapy that enables deprescription and sustained clinical improvements would be valuable. Thus, the purpose of this real-world study was to assess the effect of GLP-1 deprescription on glycemia and body weight following co-therapy with carbohydrate restricted nutrition therapy (CRNT) supported via telemedicine in a continuous remote care model among people with type 2 diabetes and excess body weight.

**Research Design and Methods:** A retrospective, propensity score matched cohort study among patients with type 2 diabetes at a nationwide telemedicine clinic was conducted using medical record data. Patients in whom GLP-1 were deprescribed (DeRx; n=154) were matched 1:1 with patients in whom GLP-1 were continued (Rx) or never prescribed (NonGLP). Longitudinal and between matched cohort differences in HbA1c and weight were assessed at enrollment, deprescription/index date, and 6 and 12 months (m) post-deprescription/index date using a linear mixed effects model.

**Results:** Hemoglobin A1c and body weight measured 6 and 12 months following deprescription/index date did not significantly differ between cohorts and improved at deprescription/index date and at follow up intervals compared to enrollment. HbA1c rose 6-and 12m post-deprescription/index in both DeRx and Rx and at 12m in NonGLP (p<0.001) but most patients maintained A1c<6.5%. No regression in body weight was observed with >70% maintaining ≥5% body weight loss 12m post-deprescription/index date.

**Conclusions:** These results demonstrate that CRNT in a continuous remote care model provides an effective GLP-1 step-off and maintenance therapy, allowing patients to discontinue GLP-1 while maintaining body weight loss and glycemia below therapeutic targets.

**KEY MESSAGES:** *What is already known on this topic:* Glucagon-like peptide-1 receptor agonists (GLP-1) have demonstrated in clinical trials significant reductions in glycemia and body weight among patients with type 2 diabetes and obesity with rapid regression of clinical improvements upon discontinuation of the medication even with persistent caloric restriction and exercise counseling, suggesting the drug must be continued indefinitely. Cost and poor persistence of the GLP-1 therapy pose real-world challenges to maintaining improved health outcomes long-term, so therapies that enable deprescription and maintenance of clinical improvements are needed.

*What this study adds:* This study assessed the potential for utilization of carbohydrate restricted nutrition therapy (CRNT) supported via telemedicine in a continuous remote care model as a GLP-1 step-off and subsequent maintenance therapy. HbA1c and body weight up to 12 months following GLP-1 deprescription did not differ from matched cohorts in whom GLP-1 were continued or never utilized in this real-world study.

*How this study might affect research, practice or policy:* This study informs clinical practice, showing that the CRNT supported by continuous remote care provides an effective GLP-1 step-off therapy, enabling maintenance of improved glycemia and weight loss following GLP-1 deprescription and mitigating the need for lifetime, continuous use of the pharmaceutical.

## INTRODUCTION

About one in seven adults in the United States lives with type 2 diabetes,[1] and 78% also live with excess weight or obesity.[2] Prevalence of type 2 diabetes, excess weight, and obesity continue to grow[3,4] alongside the cost of healthcare for these conditions,[5,6] particularly through introduction of high cost medications associated with significant weight loss, such as glucagon-like peptide-1 receptor agonists (GLP-1).[7,8]

Recent pharmaceutical advancements among incretin mimetics like GLP-1 show great potential, having elicited substantial glucose-lowering effects in type 2 diabetes[9,10] and weight loss nearing that which is achieved through surgical intervention among people with excess weight or obesity without type 2 diabetes.[11,12] However, clinical trial evidence to date demonstrates the high efficacy and high cost drugs must be continued indefinitely to sustain improved clinical outcomes.[13,14]

Lifestyle intervention, as the cornerstone of type 2 diabetes and obesity care, may serve as an effective combination and sequential therapy to pharmaceuticals to enable eventual deprescription – particularly among interventions demonstrated to elicit significant weight loss, regression of prediabetes to normoglycemia, and remission of type 2 diabetes, such as carbohydrate restricted nutrition therapy.[15,16] Using real-world data, we assessed the impact of GLP-1 deprescription on glycemia and body weight among people with type 2 diabetes and excess weight or obesity compared to matched cohorts of patients who continued GLP-1 therapy and never utilized GLP-1 therapy to evaluate the feasibility of and potential for utilization of carbohydrate restricted nutrition therapy (CRNT) supported via telemedicine in a continuous remote care model as a GLP-1 step-off therapy.

## RESEARCH DESIGN AND METHODS

### Study population and design

This retrospective real-world analysis utilized de-identified data obtained from medical records among patients of Virta Health, a nationwide telemedicine clinic serving people with type 2 diabetes, prediabetes, and obesity. Patients in the clinic are advised in carbohydrate restricted nutrition therapy, including a well-formulated and medically-supervised ketogenic diet, through use of a health application (app) where patients connect with their care team consisting of a medical provider and health coach and have continuous access to biomarker logging, educational resources, and peer support.

We identified patients with a diagnosis of type 2 diabetes who were prescribed a GLP-1 upon enrollment in the clinic whose GLP-1 was subsequently deprescribed following improved glycemia to A1c<6.5% within 3-9 months of beginning CRNT (GLP-1 deprescription cohort). To assess A1c and weight changes after deprescription in context with alternate therapeutic strategies, two matched cohorts were identified: 1) patients who were prescribed GLP-1 at enrollment and remained on GLP-1 therapy (continued GLP-1 therapy cohort) and 2) patients who were never prescribed a GLP-1 (non-GLP-1 therapy cohort).

### Outcomes and study measures

The primary aim was to determine if HbA1c or body weight differed among the GLP-1 deprescription, continued GLP-1 therapy, and non-GLP-1 therapy cohorts in the year following deprescription or index date. In addition to HbA1c, body weight, and diabetes medication data, demographics and app data, including gender, age, race and ethnicity, and daily fingerstick blood beta-hydroxybutyrate (BHB; a biomarker of adherence to the CRNT) were obtained for this analysis.

### Statistical method

To adjust for confounders and minimize bias, propensity score matching was used to match each patient in the GLP-1 deprescription cohort (reference cohort) 1:1 with a patient in the other two therapeutic cohorts. The reference cohort was matched using propensity scores estimated from a multivariate regression model and the nearest neighbor method without any replacement. For matching, enrollment and index date covariates included age, gender, race and ethnicity, HbA1c, body mass index (BMI), number of diabetes medications, and distribution of follow-up HbA1c and weight data availability and the GLP-1 drug prescribed at enrollment. To assess balance between the cohorts after matching, baseline covariates were assessed using ANOVA or Chi-Square test in all three cohorts and standardized differences were assessed between cohorts.

Longitudinal and between matched cohort differences in HbA1c and weight were assessed at enrollment, deprescription or index date, and 6 and 12 months post-deprescription or index date using linear mixed effects models. Additionally, we repeated the analyses in two medication subgroups with sufficient sample size (semaglutide and dulaglutide).

We assessed longitudinal changes in BHB in the matched cohorts using two different methods. First, the daily BHB measurements were compiled as count data where percentage days of logging BHB ≥ 0.3mM (indicative of carbohydrate restriction and low levels of nutritional ketosis) were calculated for the four main time intervals: enrollment to deprescription or index date, deprescription or index date±3 months, and 6±3 and 12±3 months post-deprescription or index date. We then used generalized estimating equations (GEE) with an unstructured correlation matrix, logarithmic link, and Poisson distribution to assess longitudinal changes and rate of change in frequency of BHB ≥ 0.3mM between the three matched cohorts. Second, mean BHB was calculated for the four main time intervals and a linear mixed effect model was used to assess longitudinal changes in mean BHB and the rate of mean BHB changes between the three matched cohorts.

All analyses were performed using R (R Foundation for Statistical Computing, Vienna, Austria) version 4.2.2 (2022-10-31) and IBM SPSS statistics (version 29.0.1.0). Two-sided *p* values less than 0.05 were considered statistically significant.

## RESULTS

Patient characteristics of the propensity-score matched cohorts are described in Table 1. No significant differences were observed between matched cohorts, and cohorts were balanced according to absolute standardized differences. Within the deprescription cohort, the medication most frequently prescribed at enrollment in the clinic was dulaglutide (43.5%), followed by semaglutide (29.2%), liraglutide (17.5%), and exenatide (7.1%). The mean duration of care in all three cohorts was at least 18 months.

**Table 1.** Baseline characteristics of the matched cohorts

|  | **GLP-1**<br>**Deprescription**<br>**Cohort (n=154)** | **Continued GLP-1**<br>**Therapy Cohort**<br>**(n=154)** | **Non-GLP-1**<br>**Therapy Cohort**<br>**(n=154)** |
| --- | --- | --- | --- |
| **Age, mean (SD), years** | 55.9 (8.7) | 55.3 (8.4) | 55.5 (9.2) |
| **Gender, n (%)** |  |  |  |
| Female | 77 (50.0) | 90 (58.4) | 84 (54.5) |
| Male | 77 (50.0) | 64 (41.6) | 70 (45.5) |
| **Race and Ethnicity (n,%)** |  |  |  |
| Non-Hispanic White | 100 (64.9) | 108 (70.1) | 105 (68.2) |
| Non-Hispanic Black or African American | 9 (5.8) | 19 (12.3) | 13 (8.4) |
| Hispanic | 29 (18.8) | 19 (12.3) | 19 (12.3) |
| Non-Hispanic American Indian or Alaska Native, Asian, Native Hawaiian or Other Pacific Islander, or Multiple Races | 7 (4.5) | 8 (5.2) | 13 (8.4) |
| **Enrollment BMI mean (SD), kg/m<sup>2</sup>** | 35.7 (6.9) | 36.5 (7.2) | 36.2 (7.8) |
| **Enrollment HbA1c mean (SD), %** | 7.3 (1.2) | 7.4 (1.4) | 7.3 (1.0) |

Hemoglobin A1c and body weight measured 6 and 12 months following deprescription or index date did not significantly differ between cohorts (Figure 1). In all cohorts, HbA1c and body weight improved significantly at time of deprescription or index date and at follow up intervals compared to enrollment. In both the deprescription and continued therapy cohorts, HbA1c at 6 and 12 month follow up rose relative to deprescription or index date (p<0.001); in the non-GLP-1 therapy cohort, HbA1c rose at 12 months post-index date (p<0.001). However, HbA1c for most individuals remained below 6.5% up to 12 months following deprescription or index date in all cohorts (deprescription: 64.8%; continued: 64.1%; non-GLP-1 therapy: 67.6%) including 20.4%, 20.3%, and 20.6% of the deprescription, continued, and non-GLP-1 therapy cohorts who maintained normoglycemia (HbA1c<5.7%) 12 months following deprescription or index date. No significant regression of body weight following deprescription or index date was observed in any cohort. More than 70% of patients in each of the matched cohorts maintained at least 5% body weight loss 12 months following deprescription or index date (Figure 2). Subgroup analyses of semaglutide and dulaglutide were consistent with the full cohort and HbA1c and body weight changes by medication are described in Figure 3.

**Figure 1.**
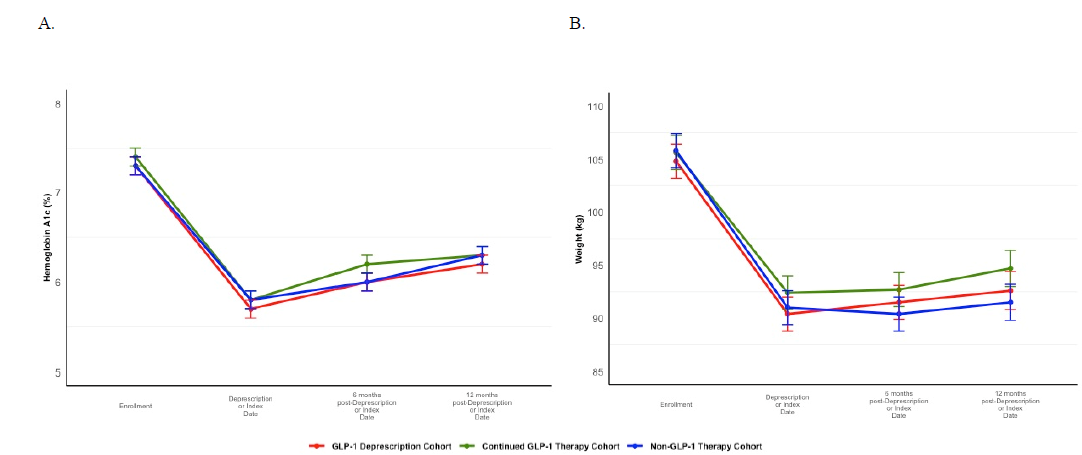
Comparison of HbA1c and body weight. Longitudinal and between group change in estimated mean (A) hemoglobin A1c (HbA1c, %) and (B) body weight (kg) from enrollment to 12 months post deprescription or index date in GLP-1 deprescription, continued GLP-1 therapy, and non-GLP-1 therapy cohorts.

**Figure 2.**
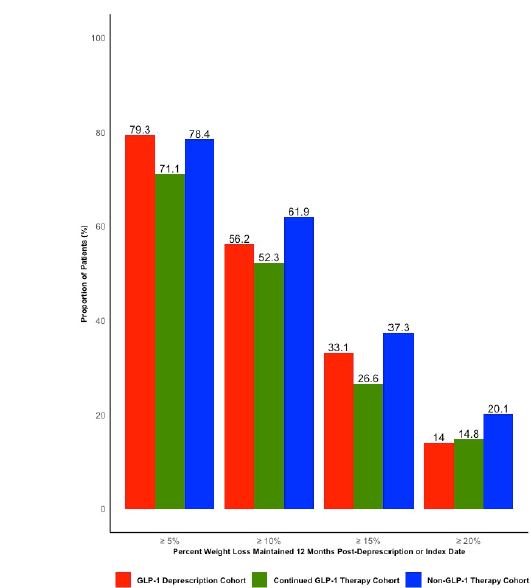
Proportion of patients maintaining weight loss targets at 12-months post-deprescription or index date by cohort.

**Figure 3.**
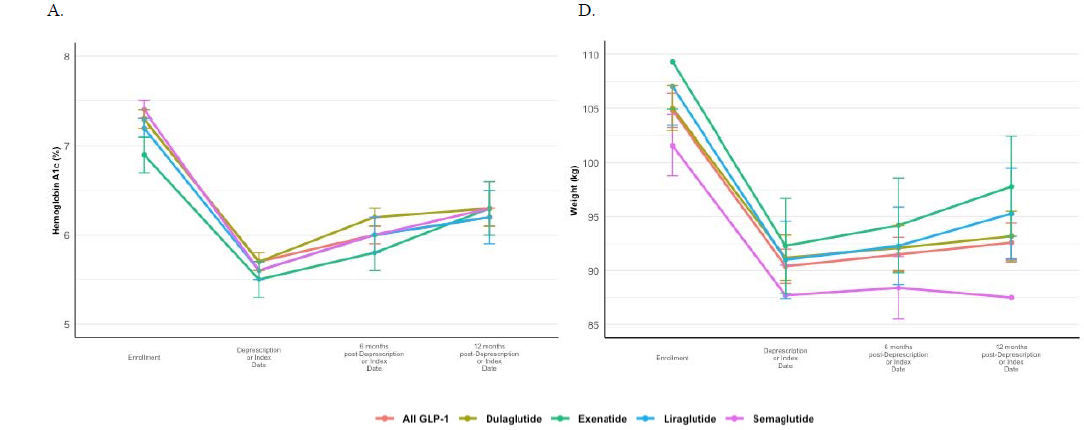
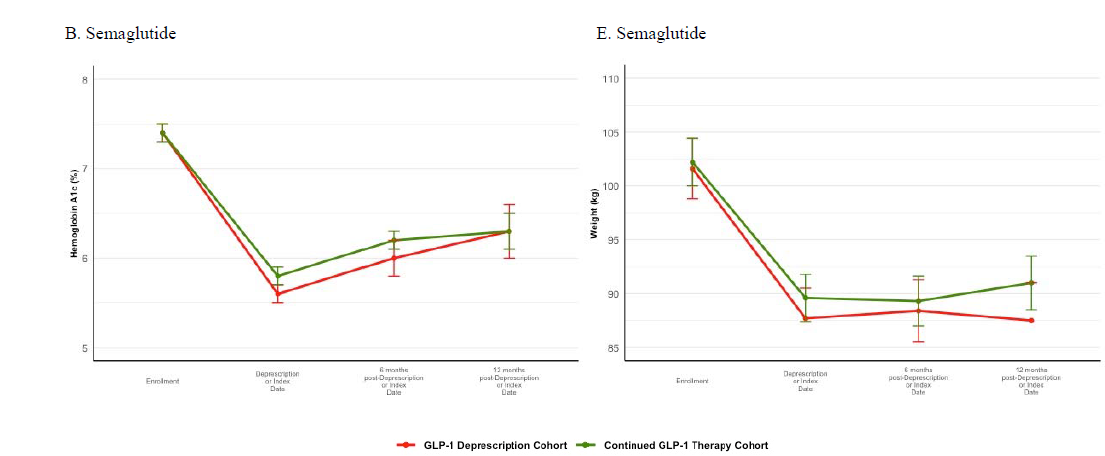
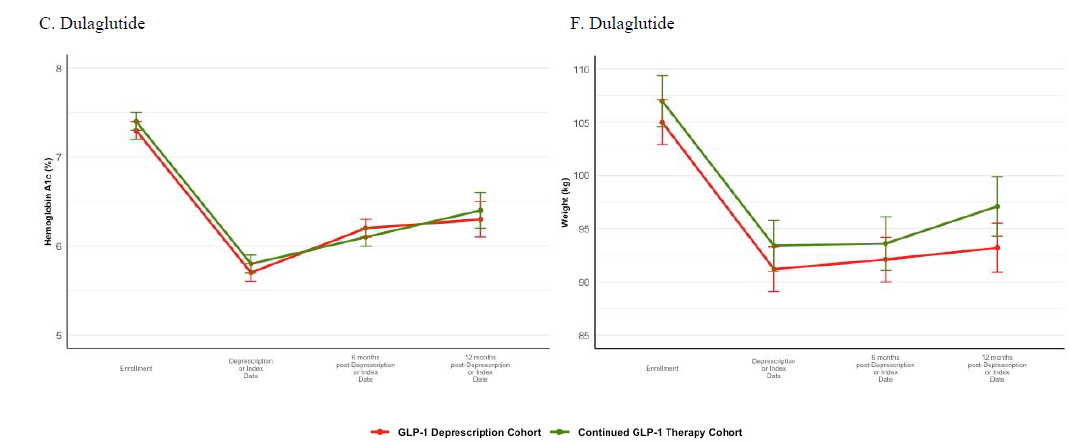
Comparison of change in HbA1c and body weight by GLP-1 medication. Longitudinal change in estimated means from enrollment through 12 months post deprescription or index date: A. HbA1c (%) in GLP-1 deprescription cohort by types of GLP-1; all GLP-1, dulaglutide, exenatide, liraglutide and semaglutide B. HbA1c (%) in subcohort analysis in those initially prescribed or continued prescribed semaglutide in GLP-1 deprescription and continued GLP-1 therapy cohorts C. HbA1c (%) in subcohort analysis in those initially prescribed or continued prescribed dulaglutide in GLP-1 deprescription and continued GLP-1 therapy cohorts D. Weight (kg) in GLP-1 deprescription cohort by types of GLP-1; all GLP-1, dulaglutide, exenatide, liraglutide and semaglutide E. Weight (kg) in subcohort analysis in those initially prescribed or continued prescribed semaglutide in GLP-1 deprescription and continued GLP-1 therapy cohorts F. Weight (kg) in subcohort analysis in those initially prescribed or continued prescribed dulaglutide in GLP-1 deprescription and continued GLP-1 therapy cohorts

Frequency of achieving BHB ≥ 0.3mM via CRNT declined more rapidly among the continued GLP-1 therapy cohort compared to the other cohorts (p=0.037), and mean BHB of the continued GLP-1 therapy cohort was lower compared to the other cohorts at all time intervals (p<0.05; Figure 4).

**Figure 4.**
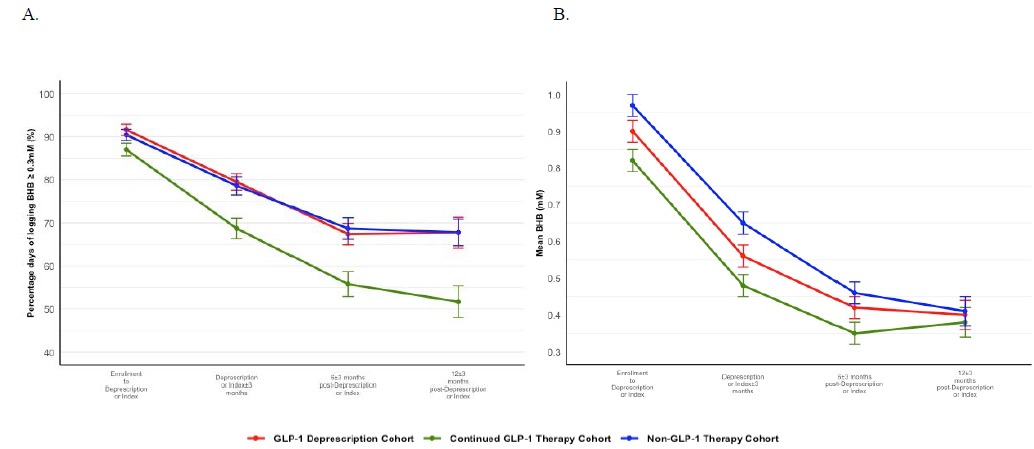
Comparison of nutritional ketosis parameters by cohort. Longitudinal change in estimated mean (A) percentage days of logging BHB ≥0.3mM and (B) BHB (mM) from enrollment to 12±3 months post deprescription or index date in GLP-1 deprescription, continued GLP-1 therapy, and non-GLP-1 therapy cohorts

## CONCLUSIONS

Results of this real-world analysis show no differences in glycemia or body weight up to 12 months following deprescription of GLP-1 in people with type 2 diabetes compared to matched cohorts in whom GLP-1 therapy was continued or no GLP-1 therapy was utilized, suggesting that carbohydrate restricted nutrition therapy supported via a continuous remote care model may be effectively utilized to step off GLP-1 therapy and used as a maintenance therapy. More frequent maintenance of nutritional ketosis achieved through CRNT was observed in the deprescription and non-GLP1 therapy cohorts during the deprescription or index time interval compared to the continued GLP1 therapy cohort, providing a potential indicator for likelihood of maintaining HbA1c and body weight following GLP-1 deprescription and tool for guiding patient care and assisting with clinical decision making regarding deprescription.

The STEP 1 Trial Extension showed rapid regression in glycemia and body weight following withdrawal of semaglutide administered in conjunction with a physical activity and caloric restriction lifestyle intervention.[13] One year after therapy withdrawal, participants regained 64% of weight lost and 80% of the decline in HbA1c that had been achieved. Although the STEP 1 Extension did not include people with type 2 diabetes, it is reasonable to expect similar regression upon withdrawal of the drug among those with type 2 diabetes. Results from the present real-world analysis contrast prior research, showing less regression of outcomes — only 15% of the body weight lost and 36% of the HbA1c decline achieved in combination with CRNT prior to GLP-1 deprescription in the year following discontinuation of the medication, despite being in a group with more progressive insulin resistance. Specifically among those deprescribed semaglutide, there was no regression in body weight one year following discontinuation and a 40% regression in the HbA1c decline. Given these patients were prescribed GLP-1 therapy prior to initiating care at this clinic, it is reasonable to expect GLP-1 therapy resulted in significant HbA1c and body weight reductions prior to those achieved by adding CRNT and continuous remote care, suggesting the overall regression in HbA1c and body weight in the context of GLP-1 deprescription and continued CRNT may be even less than what can be observed in these data.

The STEP 4 trial assessed withdrawal of the medication, but not the lifestyle intervention, and showed regain of about half of the weight lost during combination therapy over the next 11 months. The lifestyle intervention studied in the STEP 1 extension and STEP 4 trial focused on caloric restriction and exercise with monthly in-person or telephone counseling, while the lifestyle intervention in the present real-world study focuses primarily on CRNT with continuous remote support, suggesting that the type of nutrition therapy and degree of support utilized as a combination and sequential therapy may play a key role in the ability to maintain weight loss following discontinuation of the GLP-1.

One potential reason dietary carbohydrate restriction, and nutritional ketosis in particular, may provide an advantage for weight loss maintenance following GLP-1 deprescription is through reduced hunger and appetite — an effect shared by both the drug and the nutrition therapy. Participants in a clinical trial evaluating the effects of the CRNT utilized in the present study reported reduced perceptions of hunger after ten weeks of therapy concurrent with mean blood BHB of 0.6mM,[17] and blood BHB concentrations are associated with lower concentrations of the hunger hormone ghrelin and higher concentrations of satiety hormones glucagon-like peptide-1 and cholecystokinin.[18] Given the differences in BHB concentrations and frequency with which nutritional ketosis was maintained between the cohorts, blood BHB concentrations also appear to support clinical decision making regarding GLP-1 deprescription in addition to supporting patients in their daily nutrition choices and may be a useful indicator of likelihood of success in maintaining clinical improvements upon deprescription.

Longitudinal changes in HbA1c did not differ between deprescription and continued therapy cohorts, though the frequency of achieving BHB ≥ 0.3 mM with CRNT and mean BHB was higher within the deprescription cohort. Carbohydrate restriction results in less glycemic variability[19], particularly post-meal – an effect similar to that which is achieved with GLP-1 therapy through delayed gastric emptying.[20,21] Further, reduced carbohydrate intake may to some degree replace the need for glucose-dependent insulin secretion with GLP-1 therapy.

Another noteworthy observation from this study was that patients on GLP-1 therapy at enrollment achieved an additional 13% weight loss and 1.6% reduction in HbA1c upon initiating CRNT in combination with GLP-1 therapy. Further improvement in glycemia and weight elicited with this combination therapy exceeds effects observed in other real-world studies among those who switched to injectable semaglutide from less potent GLP-1.[22,23] Further, the weight loss achieved with carbohydrate restriction and GLP-1 combination therapy was on par or greater than weight loss observed in STEP 2 among people with type 2 diabetes treated with 2.4 mg and 1.0 mg semaglutide[10], suggesting there may be benefit to pairing GLP-1 with carbohydrate restricted nutrition therapy to achieve greater weight loss when clinically indicated or to enable greater weight loss when higher doses are poorly tolerated, prior to stepping off GLP-1 therapy.

Additionally, cost and side effects are important considerations in GLP-1 therapy, and may contribute to the rates of uptake, adherence, and persistence observed throughout the United States today. For example, real world persistence of GLP-1 therapy at one year is approximately 50%.[24] This suggests multiple therapeutic options must be accessible to enable the desired clinical outcomes for individual patients with unique preferences and circumstances. The achievement of significant weight loss among those who never utilized GLP-1 therapy in this analysis suggests CRNT with continuous remote care may provide a reasonable, effective, and lower cost alternative for those who contend with access, financial, or tolerability barriers or prefer nonpharmacological therapy.

Strengths of this analysis include its use of real-world data from a nationwide clinic, broadening the applicability of its findings, and that it is, to the best of the authors’ knowledge, the first study to assess glycemia and weight outcomes following withdrawal of GLP-1 in type 2 diabetes and under free-living, real-world conditions. Use of real-world data also has limitations given its retrospective and observational nature, even though differences between cohorts were reduced using matched cohort analysis. Duration of GLP-1 use prior to enrollment in the clinic and adherence could not be accounted for. Application of these findings are limited to the medications utilized by the patient population included in this analysis. Future research should evaluate the effect of GLP-1 deprescription including medications which have recently come to market as indicated for type 2 diabetes and for excess weight or obesity without type 2 diabetes.

In conclusion, results of this real-world analysis demonstrate that GLP-1 can be deprescribed without negative effects on glycemia and body weight following co-therapy with carbohydrate restricted nutrition therapy supported in a continuous remote care model compared to matched cohorts in which GLP-1 therapy was continued or not utilized. These real-world data contrast clinical trial evidence in which rapid weight regain was observed following discontinuation of GLP-1 therapy even when traditional caloric restriction and physical activity counseling persisted and suggest that carbohydrate restricted nutrition therapy and continuous care may provide an appropriate glycemia and body weight maintenance therapy, mitigating the need for lifelong, continued GLP-1 therapy.

## Data Availability

Data may be obtained from a third party and are not publicly available.

## ACKNOWLEDGMENTS

The authors would like to thank Mitch Graves, Ed Jiang, and Pang Weirich for their assistance with data compilation and Caroline Roberts, Greeshma Shetty, and Jeff Stanley for sharing their clinical insight to inform this research.

## CONTRIBUTORS

AM and SA contributed to the concept, design, and planning of the study as well as interpretation of the study results. SA analyzed the data. AM drafted the manuscript and SA reviewed.

## ETHICS APPROVAL

This study did not require IRB approval as it was a retrospective databased study in which subjects were properly de-identified to maintain confidentiality. No human subjects were at risk during this study.

## DATA AVAILABILITY STATEMENT

Data may be obtained from a third party and are not publicly available.

